# Delay in antiretroviral therapy initiation is associated with worse mood symptoms in adults with chronic HIV disease

**DOI:** 10.64898/2026.10.07.26364968

**Authors:** Hyunchan Lee, Claire O’Connor, Grace Hanly, Linxi Chen, Danial Mahmood, David J. Moore, Ronald J. Ellis, Xiong Jiang

## Abstract

**Background:** In the era of successful antiretroviral therapy (ART), mood disturbance remains highly prevalent in people with HIV (PWH). In the present study, we investigated the long-term legacy effects of long delay in ART initiation and low nadir CD4 count on mood symptoms in adults with HIV. The impacts of demographic and other HIV clinical factors were also examined.

**Methods:** A total of 109 PWH (disease duration: 24.6±9.3 years) and 42 demographically matched healthy participants (people without HIV, or PWoH) participated in a cross-sectional study (41-70 years old (55.6±6.8), 62.9% African Americans, 27.2% female, 14.3±2.9 years of education). Mood symptoms were assessed with the Beck Depression Inventory-II (BDI-II) and the Profile of Mood States (POMS). A comprehensive neuropsychological test battery was administered to assess neurocognitive function.

**Results:** Despite largely comparable neurocognitive performance between PWH and PWoH, PWH had worse mood symptoms than PWoH, including higher total BDI-II (*p*<.001) and total POMS scores (*p=*.002), as well as higher score in all POMS sub-scores (*p*<.007), except Vigor (*p*=.724). Within the PWH group, longer delays in ART initiation, lower nadir CD4 count, and lower educational attainment were associated with worse mood symptoms. By contrast, current plasma CD4+ cell count, plasma detectable viral load, and HIV disease duration were not associated with either BDI-II or POMS score.

**Conclusion:** Mood disturbance is highly prevalent in PWH on ART. The long-term legacy effects of delay in ART initiation and low nadir CD4 count on mood management reinforce the necessity of early ART initiation in clinical practice.

## INTRODUCTION

Over the past few decades, the advancement of antiretroviral therapy (ART) has led to markedly reduced mortality related to the human immunodeficiency virus (HIV) while effectively suppressing disease progression and transmission [1]. However, despite largely successful viral suppression with highly effective ART, HIV-associated brain injury persists in people with HIV (PWH), and neurocognitive impairment and mood disorders (especially depression and anxiety) remain highly prevalent. It is estimated that the prevalence of mood disorders in PWH is approximately two to three times higher than that in the general population [2–5]. Importantly, these mood symptoms do not merely develop as a consequence of HIV infection, instead they simultaneously act as critical risk factors that negatively impact both mortality and treatment adherence while exacerbating functional impairments [2,3,6].

A number of biological and psychosocial factors have been proposed to contribute to the high prevalence of mood disorders in PWH [2,7–9]. Neurotoxicity due to HIV proteins and certain antiretroviral regimens has been shown to lead to mood disturbance (including suicidal behavior) and other neurological complications [10–13]. In addition, mild neuroinflammation persists in PWH in the era of successful ART [14]. Given the critical role of neuroinflammation in depression and other neuropsychiatric disorders [15,16], HIV-related chronic neuroinflammation – despite being generally mild – may contribute to the high prevalence of depression and other mood disorders in PWH [17], supported by the evidence of associations between elevations of central and peripheral inflammation markers and worse mood states such as depression in PWH [18–20]. Less optimal viral suppression (i.e., detectable HIV viral load) and/or poor immune restoration (i.e., low CD4^+^ cell count) have also been linked to worse mood states [21–23], although the relationship is likely bidirectional and multifaceted.

Compared to the general population, people with HIV tend to have lower educational attainment and other socioeconomic statuses [24], which has been associated with greater risks of mood disorders in PWH as well as in the general population [25,26]. HIV-related stigma is a major and common psychosocial challenge constantly faced by people living with HIV [27] and has been shown to contribute to mood disorders and other mental health issues, including anxiety, depression, and suicidal ideation [28,29]. The experience of childhood trauma – a known risk factor of mood disorders [30–32] – is highly prevalent in PWH and has been associated with both physical and mental health problems [33]. Moreover, numerous studies have shown that mood disorders such as anxiety and depression often co-occur with substance use [34]; thus, the high prevalence of substance use disorders in PWH may contribute to their mood disorders [35], although the causal relationship between mood disorders and substance use disorders is complicated and remains an open question of active research [36]. These psychosocial factors can lead to undesirable neurobiological consequences contributing to mood disorders and other brain disorders. Importantly, all these biological and psychosocial factors are not mutually exclusive and often interact with one another in affecting brain health and functions in PWH, including depression and other mood disorders.

While antiretroviral therapies have been shown to lead to mood normalization [37–41] and large clinical trial studies have provided convincing evidence supporting the benefits of early ART initiation on immune restoration [42–45] and brain health [46], little is known about the long-term impact of different length of delay in ART initiation on mood symptoms and disorders (i.e., is a longer delay in ART initiation associated with worse mood disturbance?). In addition, while the status of HIV viral suppression and current CD4^+^ count have been shown to correlate with current mood symptoms, the impacts of low nadir CD4 count – reflecting a history of severe immunosuppression – on mood symptoms is not well established [47], despite its known impact on brain health and function [48–50]. In this study with middle-aged to older adults with chronic HIV disease, we aimed to investigate the legacy effects of long delay in ART initiation and low nadir CD4 count, along with the effects of other HIV and demographic factors on mood states and symptoms.

## METHODS

### Participants

A total of 109 PWH and 42 demographically matched healthy comparison participants (people without HIV, or PWoH) (41-70 years old (55.6±6.8), 62.9% Black/African Americans, 27.2% female at birth, 14.3±2.9 years of education) from the greater Washington D.C. metropolitan area participated in the study (Table 1). A telephone screen followed by an onsite screening visit was conducted to exclude participants who met one or more of the following exclusion criteria: younger than 40 or older than 70 years old; less than eight years of education; inability to speak or understand English; use of illicit substances within the past three months of the onsite screening and/or a positive urine toxicology test during the onsite screening visit; and other confounding factors such as psychosis, stroke, significant previous head trauma, and other neurological disorders unrelated to HIV. Written informed consent approved by the Institutional Review Board at Georgetown University Medical Center was obtained prior to study enrollment.

**Table 1.** Group characteristics of people with HIV (PWH) and people without HIV (PWoH) participants.

|  | *PWH (n = 109)* | *PWoH (n = 42)* | *p value* |
| --- | --- | --- | --- |
| Age (years) | 55.9 (6.7) | 54.8 (7.0) | .385 |
| Education (years) | 14.2 (3.0) | 14.4 (2.8) | .730 |
| Sex (% male) | 74.3% | 69.0% | .515 |
| Transgender (male to female) | 7.3% | 0% |  |
| Race/Ethnicity (% African-American) | 63.3% | 61.9% | .873 |
| Alcohol abuse (% participants) | 23.9% | 16.7% | .338 |
| Cocaine abuse (% participants) | 22.0% | 7.1% | **.033\*** |
| Cannabis abuse (% participants) | 12.8% | 7.1% | .401 |
| **HIV clinical characteristics (PWH only)** |  |  |  |
| HIV viral load (number of patients)<sup>a</sup> |  |  |  |
| Undetectable (<20 copies/ml) | 86 |  |  |
| Current CD4 count (cells/μl) | 637.0 [471.5, 919.5]<sup>a</sup> | | |
| Nadir CD4 (cells/μl) | 190.0 [51.5, 359.5]<sup>a</sup> | | |
| Nadir CD4 <200 cells/μl | 49.5% | | |
| Disease duration | 24.6 (9.3) |  |  |
| ART naïve | 0% |  |  |
| Delay in first ART initiation | 5.2 (6.3) |  |  |
| Current ART use (number of patients) | 106 |  |  |

MSM, Men who have sex with men.
Data are expressed as mean (SD) or median [IQR].
\* $p < .05$ ; Group comparisons for continuous variables were analyzed using Welch's *t*-tests or Wilcoxon rank-sum, and categorical variables were analyzed using chi-square test, except Cannabis use (which was analyzed using Fisher's exact test).
<sup>a</sup>Data missing for four participants.

### Experimental procedures

The Beck Depression Inventory-II (BDI-II) and the Profile of Mood States (POMS 65) were administered to assess depressive and other mood symptoms [51,52]. The BDI-II total score and the POMS total score were used in the main analysis. In addition, the six subscale scores of POMS were also examined, including Anger, Confusion, Depression, Fatigue, Tension, and Vigor. Alcohol and substance use history was screened via self-report and then diagnosed in accordance with DSM-IV criteria using the Composite International Diagnostic Interview (CIDI) (Andrews & Peters, 1998; Kessler et al., 1998). A comprehensive neuropsychological test battery was also administered to assess performance across seven neurocognitive domains that are commonly affected in PWH [50,53,54]. A previously published algorithm was utilized to calculate a global deficit score (GDS) for each participant and a validated GDS of greater than or equal to 0.5 and above indicates global neurocognitive impairment (Blackstone et al., 2012; Carey et al., 2004). Medical data and other comorbidities were also assessed.

For PWH only, the 1993 - Modified CDC Classification Worksheet was used to collect data related to HIV infection and history (e.g., date of first infection with HIV, how the HIV virus was contracted, lowest CD4 count) [55]. Nadir CD4 and estimated duration of HIV infection were self-reported. Previous studies have shown that self-reported nadir CD4 strongly correlates with actual medical records (if available) [56,57]. Four participants were unable to recall their lowest CD4 count and were therefore excluded from analyses related to nadir CD4. Blood specimens were collected to assess HIV viral load, current CD4+ T cell count, and current CD8 cell count.

In addition, while the first antiretroviral medicine (Zedovudine) was approved by FDA in 1987 and the dual therapy was introduced around 1992, the highly effective combination antiretroviral therapy with triple drug combination only became available around 1996 [58]. To control for potential difference in the magnitude of benefits of different ARTs (e.g., monotherapy with Zedovudine vs highly effective combination antiretroviral therapy), a categorical variable (IScART) was created and included in all data analysis, with IScART=0 for those who started antiretroviral therapy before 1996 and 1 for those who started antiretroviral therapy in or after 1996. Additional sensitivity analyses were also conducted with each of the two subgroups of PWH (i.e., IScART = 0 vs 1) separately.

### Statistical analysis

Determined by the characteristics of the corresponding data, Welch’s two-sample *t*-test, Wilcoxon rank-sum test, chi-square or Fisher’s exact test were used to examine group differences between PWH and PWoH. Bivariate relationships were assessed using Pearson correlation analysis with 5,000 bootstrap samples. Multivariable linear regression analyses were conducted among PWH to examine the associations of disease-related variables with mood symptoms while adjusting for demographic covariates. Separate regression models were fitted for BDI-II and POMS. False discovery rate (FDR) corrections were implemented in multiple comparisons and both uncorrected *p* values and corrected *q* values are reported when FDR-correction was applied.

All statistical analyses and data visualization were performed using R (R Foundation for Statistical Computing), Prism (version 11, GraphPad Software) and SPSS (version 31, IBM). R packages used for statistical analysis and visualization included dplyr, ggplot2, broom, car, sandwich, and lmtest. Data were presented as mean with standard deviation (SD) or median with interquartile range (IQR) provided in parentheses where appropriate.

## RESULTS

### Demographic and HIV clinical data

The PWH and PWoH participants were well-matched in demographics, and there were no significant group differences between PWH and PWoH, except a higher prevalence of cocaine abuse in the PWH group (*p*=.033, Table 1).

The self-reported HIV infection dates (in years) are shown in Fig. 1A, with the earliest HIV infection dated back to 1975 for one participant. The duration of disease ranged from 2 to 44 years, with a mean of 24.6±9.3 years (Fig. 1B and Table 1). The first ART initiation among this cohort of PWH participants was dated back to 1985 (Fig. 1C). The delay between first ART initiation and HIV infection ranged from 0 to 40 years, with an average delay of 5.2±6.3 years (Fig. 1D and Table 1). There was a 10-year gap between the earliest date of HIV infection and the earliest date of first ART initiation among PWH participants, suggesting that PWH infected in earlier years not only had a longer disease duration but also faced extended delays in receiving ART. Consistent with this pattern, the delay in first ART initiation was negatively correlated with disease duration (*p*<.001, Fig. 1E). Several factors likely contribute to the longer delay in ART initiation for PWH infected in earlier years: early in the HIV pandemic, antiretroviral treatment was not always covered by medical insurance thus might be out of reach for some PWH due to affordability; while the current policy recommends initiating ART right after seroconversion, some versions of early policy recommended not to initiate ART unless disease progressed to a certain point, being determined by factors such plasma CD4+ cell counts.

**Figure 1.**
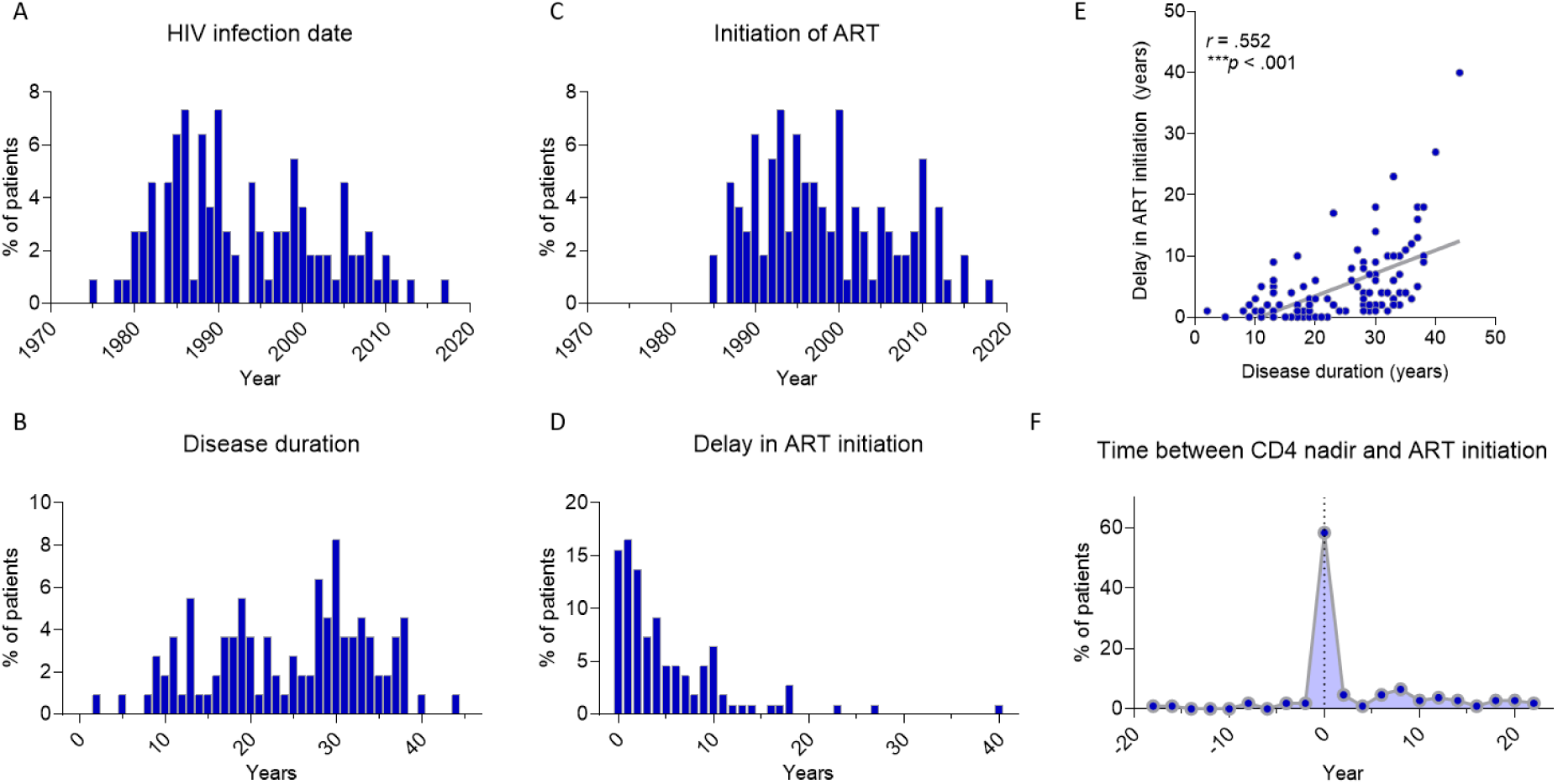
Historical trends in first HIV infection and delays in first ART initiation. (**A**) Histogram shows the proportion of PWH who have their first HIV infection each year (*n* = 109). (**B**) The distribution of disease duration among PWH (*n* = 109). (**C**) The proportion of PWH who initiate their first ART each year (*n* = 109). (**D**) The distribution of delays in their first ART initiation (*n* = 109). (**E**) Delay in first ART initiation was significantly correlated with disease duration in correlation analysis (*r* = .552, \*\*\**p* < .001, *n* = 109). (**F**) The proportion of PWH according to the time difference between nadir CD4 and first ART initiation, indicating whether nadir CD4 was recorded before, on, or after first ART initiation (*n* = 108).

### Mood symptoms and neuropsychological performance

Compared to PWoH, PWH had significantly worse depressive and other mood disturbance symptoms, including higher total BDI-II and total POMS scores, as well as higher score in all POMS sub-scores (*p*s<.007), except Vigor (*p*=.724) (Table 2). By contrast, there were no significant differences in any of the global neurocognitive measures as well as in any of the neurocognitive domains (*p*s>.209), except a marginally significant difference between PWH and PWoH in attention/working memory (*p*=.038), with PWH performing worse than PWoH, but the difference in attention/working memory did not survive correction for multiple comparisons (*q*=.281). In line with these findings, comparisons across the four area under curves measures demonstrated that the difference between PWH and PWoH was exclusively significant and more sensitively differentiated in total POMS and BDI-II scores than in any other neurocognitive measures (Supplementary Fig. S1). Furthermore, among PWH, there was no significant correlation between neurocognitive measures and total POMS or BDI-II scores (Supplementary Fig. S2A-B).

**Table 2.** Mood disturbance and neurocognitive performance.

| *Mood disturbance* | *PWH (n = 109)* | *PWoH (n = 42)* | *p value* | *q value* |
| --- | --- | --- | --- | --- |
| Profile of mood states (total) | 15.20 (29.38)<sup>b</sup><br>11.00 [-7.00, 28.00] | -1.07 (19.60)<sup>a</sup><br>-2.00 [-17.50, 12.00] | **.002\*\*** |  |
| Depression | 7.33 (7.90)<sup>a</sup><br>5.00 [2.00, 10.00] | 3.81 (4.40)<br>2.00 [0.00, 7.00] | **.006\*\*** | **.001\*\*** |
| Tension | 7.95 (6.06)<sup>a</sup><br>6.50 [3.00, 12.00] | 4.68 (3.93)<sup>a</sup><br>4.00 [1.00, 8.00] | **.002\*\*** | **<.001\*\*\*** |
| Anger | 6.56 (6.20)<sup>a</sup><br>5.00 [2.00, 9.75] | 3.38 (4.42)<br>1.00 [0.00, 5.25] | **<.001\*\*\*** | **<.001\*\*\*** |
| Fatigue | 7.52 (5.48)<br>6.00 [4.00, 10.00] | 3.60 (3.41)<br>3.00 [0.75, 6.00] | **<.001\*\*\*** | **<.001\*\*\*** |
| Confusion | 6.02 (4.32)<br>5.00 [3.00, 8.00] | 3.55 (3.14)<br>3.00 [1.00, 5.25] | **<.001\*\*\*** | **<.001\*\*\*** |
| Vigor | 20.22 (6.66)<br>20.00 [16.00, 25.00] | 20.26 (5.98)<br>19.50 [15.75, 24.25] | .724 | .122 |
| Beck's Depression Inventory | 7.96 (0.67)<br>6.00 [3.00, 11.00] | 3.83 (4.65)<br>3.50 [0.00, 6.00] | **<.001\*\*\*** |  |
| *Neurocognitive performance* | *PWH (n = 109)* | *PWoH (n = 42)* | *p value* | *q value* |
| Cognitively impaired, n(%) | 33 (30.3%) | 11 (26.2%) | .621 |  |
| Global Mean T | 48.51 (6.39) | 49.73 (7.49) | .353 |  |
| Attention/Working memory | 47.26 (9.18) | 51.67 (12.22) | **.038\*** | .281 |
| Executive function | 49.71 (10.33) | 52.12 (10.56) | .210 | .724 |
| Information processing speed | 50.99 (8.12) | 51.87 (8.449) | .563 | .827 |
| Language | 52.14 (8.22) | 53.56 (9.69) | .403 | .741 |
| Learning | 43.90 (8.37) | 44.07 (11.08) | .930 | .976 |
| Memory | 45.22 (8.84) | 44.62 (10.68) | .749 | .917 |
| Motor skills | 47.17 (9.19)<sup>b</sup> | 49.12 (10.52) | .296 | .724 |

Data are expressed as mean (SD), median (IQR), or n(%).
\* $p/q < .05$ , \*\*\* $p/q < .001$ ; Continuous variables were analyzed using Wilcoxon rank-sum (BDI-II and POMS) or Welch's $t$ -tests (neurocognitive performance), and a categorical variable (% participants $\geq 10$ ) was analyzed using chi-square test.
<sup>a</sup>Data missing for one participant.
<sup>b</sup>Data missing for two participants.

### The associations between mood symptoms and HIV clinical and demographic data in PWH

Worse current mood symptoms, including higher total BDI-II score and total POMS score, were associated with longer delay in ART initiation (*p*=.016 and .002, respectively, Fig. 2A & 2B, Table 3), but not with disease duration (*p*=.242 and .132, respectively, Fig. 2E & 2F, Table 3) or the duration of antiretroviral therapy (*p*=.480 and .257, respectively, Supplementary Fig. S3). Longer delay in ART initiation was also associated with all POMS subscale scores (*p*s<.032), except Vigor (*p*=.422) (Supplementary Materials Table S1).

**Figure 2.**
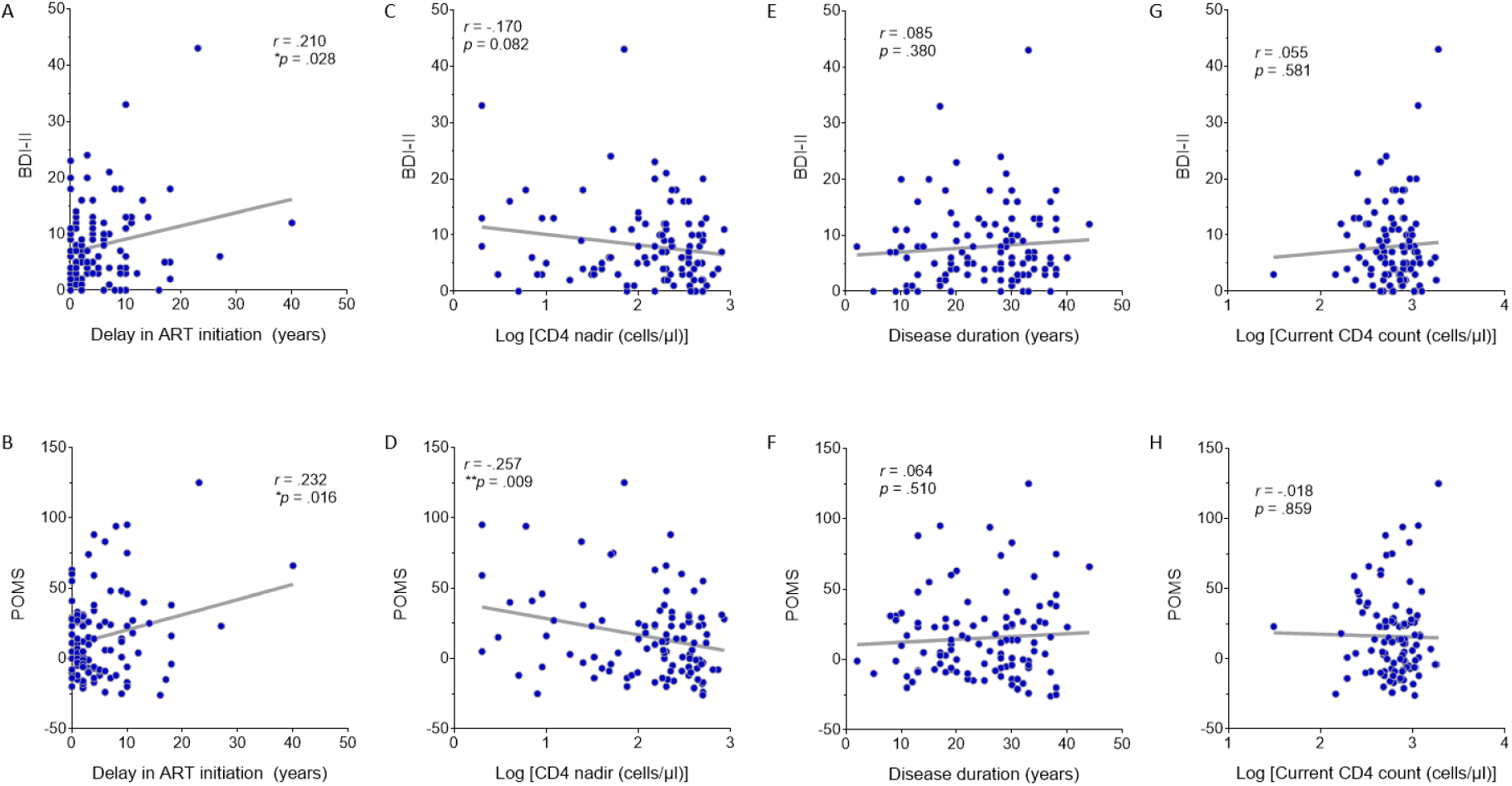
Correlation between delay in first ART initiation and mood symptoms in PWH. (**A-B**) Delay in first ART initiation was significantly correlated with total BDI-II scores (*r* = .210, \**p* = .028, *n* = 109), and with total POMS scores (*r* = .232, \**p* = .016, *n* = 107) in correlation analysis. (**C-D**) Nadir CD4 in PWH was not correlated with BDI-II (*r* = -.170, *p* = .082, *n* = 105), but was significantly correlated with total POMS scores (*r* = -.257, \*\**p* = .009, *n* = 103). (**E-F**) Disease duration showed no significant correlations with total BDI-II scores (*r* = .085, *p* = .380, *n* = 109) and total POMS scores (*r* = .064, *p* = .510, *n* = 107). (**G-H**) Current CD4 count in PWH was not significantly correlated with total BDI-II scores (*r* = .055, *p* = .581, *n* = 105) or total mood disturbance scores (*r* = -.018, *p* = .859, *n* = 103). Note: All *r* and *p* values in Fig, 2 were from bivariate analyses with Pearson’s correlation using 5,000 bootstrap samples.

**Table 3.**
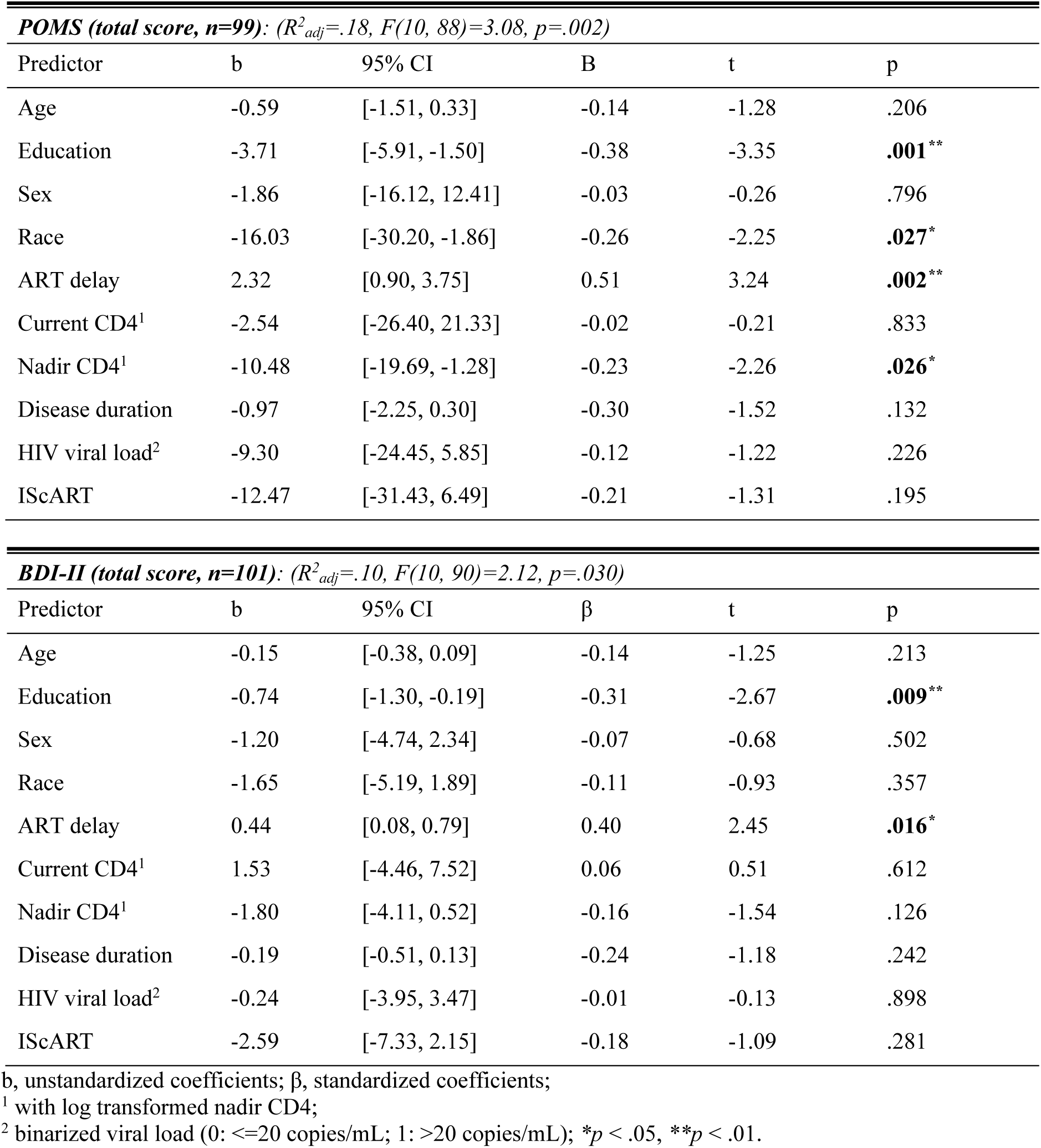
Multivariable linear regression analysis of factors associated with mood disturbance and neurocognitive impairment in people living with HIV.

Nadir CD4 counts (log-transformed) were also associated with higher total POMS score (*p*=.026) but not total BDI-II score (*p*=.126, Table 3), and there was no significant correlation between current CD4 (log-transformed) counts or HIV viral suppression (binarized as 0/1) with either total POMS (*p*=.226, Table 3) or BDI-II (*p*=.612, Fig. 2G & 2H, Table 3).

Greater educational attainment (i.e., more years of formal education) was associated with less mood symptoms, including lower total BDI-II score and total POMS score (*p*=.009 and .001, respectively). Black/African American participants had lower total POMS score (*p*=.027) but not BDI-II (*p*=.357). There was no significant association between mood symptoms (either BDI-II or POMS score) with any of the other demographic variables (Table 3).

Additional sensitivity analyses were conducted with each of the two subgroups of PWH (starting ART before vs after 1996) separately using the same statistical model. In the subgroup of PWH who started ART in or after 1996, a longer delay in ART initiation was associated with worse mood symptoms, with a significant association with total POMS score (*p*=.014) and a marginal but non-significant association with total BDI-II score (*p*=.098, Supplementary Table S1). By contrast, in the subgroup of PWH who started ART before 1996, there was no association between the length of delay in ART initiation and mood symptoms (BDI, *p*=.138; POMS, *p*=.340; Supplementary Table S2). An approximately 50% reduction in sample size likely contributed to reduced significance levels.

## DISCUSSION

In this study with a cohort of middle-aged to older adults with chronic HIV disease (disease duration 24.6±9.3 years, range 2 to 44 years) who had largely successful HIV viral suppression and immune restoration, we found that PWH had worse mood symptoms (including depressive symptoms) than demographically matched PWoH, despite largely comparable neurocognitive performance. Importantly, a longer delay in ART initiation, a lower nadir CD4 count, and lower educational attainment were associated with worse mood symptoms in PWH.

Mood disorders (especially depression and anxiety) are highly prevalent in people living with HIV, including during the acute/primary HIV infection stage [40]. Despite conflicting reports regarding the relationship between the likelihood of ART initiation and mood states (especially depression) [41,59], studies have provided largely converging evidence supporting the benefits of antiretroviral therapy in normalizing mood states [37–41] (but also see [60]). Several studies have even found that longer duration of antiretroviral therapy is associated with better mood symptoms (i.e., less than two years versus two years or longer on ART) [38,39], further supporting the benefits of antiretroviral therapy on improving mood states in PWH. In the present study, we did not find an association between mood symptoms and duration of antiretroviral therapy (*p*s>.256). By contrast, a longer delay in ART initiation was significantly associated with worse mood symptoms in PWH (BDI-II, *p*=.016; POMS, *p*=.002). Several factors may contribute to the differences: in the previous studies, the benefits of longer ART duration on mood symptoms is mainly driven by the worse mood symptoms in PWH who were on ART for less than two years [38,39], thus the effect of ART duration might be difficult to detect in the present study as PWH had a rather long disease duration (24.6±9.3 years) and their ART durations were four years or longer (except one PWH who had a disease duration of two years and was on ART for more than one year); furthermore, the association between mood symptoms and duration of delay in ART initiation might be present but were not examined in these previous studies [38,39]. The highly effective combination antiretroviral therapy with triple drug combination only became available around 1996 [58]; thus, the magnitude of benefits of early ART initiation may be different between PWH who started ART before 1996 and those who started ART after 1996. In this study, in addition to including the potential impact of different ART types (i.e., monotherapy vs combination antiretroviral therapy) as a covariate in all analyses, we also conducted additional sensitivity analyses by dividing PWH into two subgroups (depending on whether they started antiretroviral therapy before or after 1996). The additional sensitivity analyses further support an association between a longer delay in ART initiation and worse mood symptoms, despite reduced significance levels due to an approximately 50% reduction in sample size for each of the two sensitivity analyses. Interestingly, the association between the length of delay in ART initiation and mood symptoms might be stronger in PWH who started ART in or after 1996 than those who started ART before 1996, suggesting that combination antiretroviral therapy with triple drug combination may be more effective in restoring/maintaining brain health (including mood states) than the monotherapy or dual therapy available before 1996, in line with the literature [61,62]. Taken together, the significant association between delay in ART initiation and mood symptoms in PWH with chronic HIV disease is present and strong in this cohort of PWH with chronic HIV disease, suggesting a potentially long-lasting adverse effect of delay in ART initiation on mood management, with a longer delay in ART initiation associated with worse outcomes. These results reinforce the necessity of early ART initiation in clinical practice and provide additional evidence supporting the current policy of the United Nations and governments around the world.

Nadir CD4 count is the lowest ever known CD4+ count. A low nadir CD4 count, which represents a history of severe immunosuppression, has been shown to have long-lasting effects on brain structure and function [48–50,63–65]. However, despite the known impact on brain health, the evidence of low nadir CD4 on mood disturbance is scarce. Very few studies have reported an association between mood disturbance and nadir CD4 count. Kinyanda and colleagues found that both low educational attainment and low nadir CD4 count predicted incident major depressive disorder in youth living with HIV in Uganda [47]. Kapetanovic and colleagues reported that mothers with a nadir CD4 of 200 cells/mm3 or lower during pregnancy were three times more likely to experience maternal perinatal depression than mothers with a nadir CD4 of 500 cells/mm3 or higher during pregnancy [66]. In this study, with a cohort of middle-aged to older PWH with chronic HIV disease, we found that lower nadir CD4 was associated with worse POMS score (*p*=.026), suggesting a potential long-lasting legacy effect of the depth of previous immunosuppression on current mood states, which is also supported by a marginal but non-significant association between nadir CD4 count and BDI-II score (*p*=.126). By contrast, in this cohort of middle-aged to older PWH with chronic HIV disease, current mood symptoms were not associated with current CD4+ count, or disease duration, or HIV viral load (binarized as 0/1) (*p*s>.225). The relatively good health conditions among this cohort of PWH may contribute to the null findings with current CD4+ count and HIV viral load (Table 1).

Lower socioeconomic status (including educational attainment, i.e., the number of years of formal education) has been shown to be risk factor for mood disorders, including depression and anxiety [25,67,68]. Similar to the HIV-uninfected general population, lower educational attainment in PWH has been linked to worse mood states and symptoms [26,69,70]. In the present study, we found that lower educational attainment was associated with worse mood symptoms (BDI-II, *p*=.009; POMS, *p*=.001), in line with these previous studies. Interestingly, while brain imaging studies have found that mood disorders related alterations in brain structure and function in PWH are largely consistent with the findings from the general population [19,53,71,72], in a very recent study, we have found that lower educational attainment was associated with smaller basal ganglia and diencephalon in PWH, along with a weak but non-significant trend that worse depressive symptoms were associated with smaller basal ganglia and diencephalon (*p*s<.12) [72]. Given the important role of basal ganglia and diencephalon in mood disorders (including anxiety and depression) [73–76], future studies with a large sample may help to uncover the potential three-way interaction among educational attainment, mood symptoms, and the basal ganglia and diencephalon gray matter volume.

The present study has several limitations. First, this is a cross-sectional study without longitudinal data and the mood symptoms data were collected from a single study visit, thus the impacts of delay in ART initiation on mood symptoms and disorders need to be examined and verified with data from longitudinal studies, especially those with a large cohort of participants (compared to the modest sample size in the present study). Second, there are known sex differences in the behavioral profiles of mood disorders as well as the underlying neural mechanisms [77–79]. In the present study, we did not observe a significant effect of sex on mood symptoms (*p*s>.501), but the lack of significance could be due to an imbalanced sample (Table 1). Third, while both the BDI-II and the POMS are widely-used instruments in both clinical practice and basic research, neither is designed as a diagnostic tool. Therefore, the results in the present study should be interpreted with caution and should be confirmed in future studies with clinical diagnosis data from certified professionals, which was not available or feasible in the present study.

## CONCLUSIONS

Despite successful antiretroviral therapy, mood disturbance remains highly prevalent in PWH. A longer delay in ART initiation, a history of severe immunosuppression, and lower educational attainment are associated with worse mood disturbance.

## Data and Code Availability

To protect patients’ privacy, deidentified data are available upon reasonable request.

## Author Contributions

**H.L.**: Writing – first draft, Writing – review & editing, Methodology, Formal analysis, Conceptualization. **C.O.**: Writing – review & editing, Project administration. **G.H.**: Writing – review & editing, Project administration. **L.C**: Writing – review & editing, Methodology, Formal analysis. **D.M.**: Writing – review & editing, Investigation. **D.J.M.**: Writing – review & editing, Funding acquisition, Data curation, Investigation. **R.J.E.**: Writing – review & editing, Funding acquisition, Data curation, Investigation. **X.J.**: Writing – first draft, Writing – review & editing, Methodology, Funding acquisition, Data curation, Investigation, Conceptualization.

## Ethics approval and consent statement

The participants provided their written informed consent prior to participate in this study.

## Acknowledgments

We wish to thank all participants for their time and participation, and the assistance for patient care from the Georgetown University Clinical Research Unit (GU-CRU), which has been supported by Grant # UM1TR005465 (previously UL1TR001409) through the Clinical and Translational Science Awards Program (CTSA).

