## Supplementary Table S1, Supplementary Table S2, Supplementary Table S3, Supplementary Figure S1, Supplementary Figure S2, Supplementary Figure S3 for "Delay in antiretroviral therapy initiation is associated with worse mood symptoms in adults with chronic HIV disease"

### Supplementary Materials

**Table S1. Multivariable linear regression analysis of factors associated with POMS sub-scores in people living with HIV 2**

**Table S2. Multivariable linear regression analysis of factors associated with mood disturbance in people living with HIV starting ART after 1996 5**

**Table S3. Multivariable linear regression analysis of factors associated with mood disturbance in people living with HIV starting ART before 1996 6**

**Figure S1. ROC curves for mood and neurocognitive measures in PWH 7**

**Figure S2. Correlation between mood and neurocognitive measures in PWH 8**

**Figure S3. Correlation between duration of ART and mood symptoms in PWH 9**

**Supplementary Table S1. Multivariable linear regression analysis of factors associated with POMS subscale scores in people with HIV**

| ***Depression (total score, n=100)**: ($R^2_{adj}$=.19, F(10, 89)=3.27, p=.001)* | | | | | |
|---|---|---|---|---|---|
| Predictor | b | 95% CI | β | t | p |
| Age | -0.16 | [-0.40, 0.09] | -0.13 | -1.27 | .208 |
| Education | -0.69 | [-1.28, -0.09] | -0.26 | -2.30 | **.024**[*] |
| Sex | -0.22 | [-4.02, 3.58] | -0.01 | -0.11 | .909 |
| Race | -1.54 | [-5.35, 2.27] | -0.09 | -0.80 | .424 |
| ART delay | 0.61 | [0.23, 1.00] | 0.49 | 3.17 | **.002**[**] |
| Current CD4[1] | 0.30 | [-6.11, 6.71] | 0.01 | 0.09 | .926 |
| Nadir CD4[1] | -4.13 | [-6.60, -1.65] | -0.33 | -3.31 | **.001**[**] |
| Disease duration | -0.27 | [-0.62, 0.07] | -0.31 | -1.59 | .115 |
| Viral load[2] | 2.17 | [-1.90, 6.24] | 0.10 | 1.06 | .292 |
| IScART | -4.63 | [-9.71, 0.44] | -0.28 | -1.81 | .073 |

| ***Tension (total score, n=100)**: ($R^2_{adj}$=.13, F(10, 89)=2.43, p=.013)* | | | | | |
|---|---|---|---|---|---|
| Predictor | b | 95% CI | β | t | p |
| Age | -0.09 | [-0.28, 0.11] | -0.10 | -0.89 | .377 |
| Education | -0.65 | [-1.12, -0.18] | -0.32 | -2.74 | **.007**[**] |
| Sex | -0.36 | [-3.38, 2.66] | -0.03 | -0.24 | .814 |
| Race | -2.72 | [-5.74, 0.30] | -0.22 | -1.79 | .077 |
| ART delay | 0.51 | [0.21, 0.81] | 0.53 | 3.33 | **.001**[**] |
| Current CD4[1] | 0.37 | [-4.72, 5.46] | 0.02 | 0.14 | .886 |
| Nadir CD4[1] | -1.43 | [-3.39, 0.53] | -0.15 | -1.45 | .152 |
| Disease duration | -0.19 | [-0.46, 0.09] | -0.28 | -1.36 | .178 |
| Viral load[2] | 1.99 | [-1.24, 5.21] | 0.12 | 1.22 | .224 |
| IScART | -2.56 | [-6.59, 1.47] | -0.20 | -1.26 | .210 |

| ***Anger (total score, n=100)**: ($R^2_{adj}$=.12, F(10, 89)=2.30, p=.019)* | | | | | |
|---|---|---|---|---|---|
| Predictor | b | 95% CI | B | t | p |
| Age | -0.15 | [-0.35, 0.05] | -0.17 | -1.54 | .128 |
| Education | -0.69 | [-1.17, -0.21] | -0.33 | -2.85 | **.005**[**] |
| Sex | -0.27 | [-3.37, 2.82] | -0.02 | -0.18 | .861 |
| Race | -2.12 | [-5.20, 0.95] | -0.17 | -1.37 | .173 |
| ART delay | 0.42 | [0.11, 0.73] | 0.43 | 2.69 | **.009**[**] |
| Current CD4[1] | 0.04 | [-5.17, 5.24] | 0.00 | 0.01 | .989 |
| Nadir CD4[1] | -1.25 | [-3.25, 0.76] | -0.13 | -1.23 | .221 |

| | | | | | |
|---|---|---|---|---|---|
| Disease duration | -0.16 | [-0.44, 0.12] | -0.23 | -1.14 | .259 |
| Viral load[2] | 2.19 | [-1.03, 5.42] | 0.14 | 1.35 | .180 |
| IScART | -1.67 | [-5.81, 2.46] | -0.13 | -0.80 | .423 |

***Fatigue (total score, n=101)**: ($R^2_{adj}$=.09, F(10, 90)=2.04, p=.038)*

| Predictor | b | 95% CI | β | t | p |
|---|---|---|---|---|---|
| Age | -0.05 | [-0.23, 0.13] | -0.07 | -0.59 | .555 |
| Education | -0.68 | [-1.11, -0.25] | -0.37 | -3.14 | **.002**** |
| Sex | -0.15 | [-2.91, 2.60] | -0.01 | -0.11 | .912 |
| Race | -4.17 | [-6.92, -1.41] | -0.36 | -3.00 | **.003**** |
| ART delay | 0.31 | [0.03, 0.58] | 0.35 | 2.19 | **.031*** |
| Current CD4[1] | -0.11 | [-4.77, 4.56] | 0.00 | -0.04 | .964 |
| Nadir CD4[1] | -0.88 | [-2.68, 0.92] | -0.10 | -0.97 | .335 |
| Disease duration | -0.06 | [-0.31, 0.18] | -0.11 | -0.52 | .607 |
| Viral load[2] | 0.51 | [-2.38, 3.41] | 0.04 | 0.35 | .725 |
| IScART | -0.62 | [-4.32, 3.07] | -0.06 | -0.33 | .739 |

***Confusion (total score, n=101)**: ($R^2_{adj}$=.18, F(10, 90)=3.24, p=.001)*

| Predictor | b | 95% CI | β | t | p |
|---|---|---|---|---|---|
| Age | -0.02 | [-0.15, 0.12] | -0.03 | -0.29 | .773 |
| Education | -0.55 | [-0.88, -0.23] | -0.38 | -3.40 | .001 |
| Sex | 0.12 | [-1.95, 2.20] | 0.01 | 0.12 | .906 |
| Race | -2.62 | [-4.69, -0.54] | -0.29 | -2.50 | **.014*** |
| ART delay | 0.34 | [0.13, 0.55] | 0.50 | 3.24 | **.002**** |
| Current CD4[1] | 0.86 | [-2.66, 4.37] | 0.05 | 0.48 | .629 |
| Nadir CD4[1] | -2.08 | [-3.44, -0.72] | -0.31 | -3.04 | **.003**** |
| Disease duration | -0.21 | [-0.40, -0.02] | -0.44 | -2.25 | **.027*** |
| Viral load[2] | 0.91 | [-1.27, 3.09] | 0.08 | 0.83 | .407 |
| IScART | -1.88 | [-4.66, 0.90] | -0.21 | -1.34 | .183 |

***Vigor (total score, n=101)**: ($R^2_{adj}$=-.01, F(10, 90)=.90, p=.540)*

| Predictor | b | 95% CI | β | t | p |
|---|---|---|---|---|---|
| Age | 0.11 | [-0.12, 0.34] | 0.11 | 0.98 | .332 |
| Education | 0.47 | [-0.09, 1.02] | 0.21 | 1.68 | .097 |
| Sex | 1.47 | [-2.06, 4.99] | 0.10 | 0.83 | .410 |
| Race | 2.85 | [-0.67, 6.37] | 0.21 | 1.61 | .112 |
| ART delay | -0.14 | [-0.50, 0.21] | -0.14 | -0.81 | .422 |

| | | | | | |
|---|---|---|---|---|---|
| Current CD4[1] | 3.89 | [-2.08, 9.85] | 0.15 | 1.30 | .199 |
| Nadir CD4[1] | 0.74 | [-1.56, 3.04] | 0.07 | 0.64 | .523 |
| Disease duration | 0.09 | [-0.22, 0.41] | 0.13 | 0.59 | .556 |
| Viral load[2] | -1.89 | [-5.58, 1.81] | -0.11 | -1.01 | .313 |
| IScART | 1.49 | [-3.23, 6.21] | 0.11 | 0.63 | .533 |

b, unstandardized coefficients; β, standardized coefficients;
[1] with log transformed nadir CD4;
[2] binarized viral load (0: <=20 copies/mL; 1: >20 copies/mL); $*p < .05$, $**p < .01$.

**Supplementary Table S2. Multivariable linear regression analysis of factors associated with mood disturbance in people living with HIV starting ART after 1996**

| ***POMS (total score, n=59)***: *($R^2_{adj}$=.17, F(9, 49)=2.31, p=.030)* | | | | | |
|---|---|---|---|---|---|
| Predictor | b | 95% CI | B | t | p |
| Age | -1.08 | [-2.37, 0.21] | -0.23 | -1.68 | .100 |
| Education | -4.26 | [-7.31, -1.22] | -0.43 | -2.81 | **.007**** |
| Sex | 7.74 | [-13.02, 28.51] | 0.11 | 0.75 | .457 |
| Race | -4.74 | [-25.84, 16.36] | -0.08 | -0.45 | .653 |
| ART delay | 2.23 | [0.47, 3.99] | 0.57 | 2.55 | **.014*** |
| Current CD4[1] | 6.45 | [-23.88, 36.79] | 0.06 | 0.43 | .671 |
| Nadir CD4[1] | -6.08 | [-19.11, 6.95] | -0.13 | -0.94 | .353 |
| Disease duration | -1.11 | [-2.71, 0.49] | -0.33 | -1.40 | .168 |
| Viral load[2] | 6.71 | [-12.59, 26.01] | 0.09 | 0.70 | .488 |

| ***BDI-II (total score, n=59)***: *($R^2_{adj}$=.19, F(9, 49)=2.48, p=.020)* | | | | | |
|---|---|---|---|---|---|
| Predictor | b | 95% CI | B | t | p |
| Age | -0.32 | [-0.66, 0.03] | -0.25 | -1.86 | .068 |
| Education | -0.70 | [-1.50, 0.11] | -0.26 | -1.74 | .087 |
| Sex | 0.70 | [-4.77, 6.18] | 0.04 | 0.26 | .797 |
| Race | 2.25 | [-3.31, 7.81] | 0.14 | 0.81 | .420 |
| ART delay | 0.39 | [-0.08, 0.85] | 0.37 | 1.69 | .098 |
| Current CD4[1] | 4.80 | [-3.19, 12.80] | 0.16 | 1.21 | .233 |
| Nadir CD4[1] | -1.57 | [-5.00, 1.86] | -0.12 | -0.92 | .362 |
| Disease duration | -0.21 | [-0.63, 0.21] | -0.23 | -0.99 | .328 |
| Viral load[2] | 4.71 | [-0.37, 9.80] | 0.23 | 1.86 | .069 |

b, unstandardized coefficients; β, standardized coefficients;
[1] with log transformed nadir CD4;
[2] binarized viral load (0: <=20 copies/mL; 1: >20 copies/mL); **p* < .05, ***p* < .01.

**Supplementary Table S3. Multivariable linear regression analysis of factors associated with mood disturbance in people living with HIV starting ART before 1996**

| ***POMS (total score, n=38)***: *($R^2_{adj}$=.14, F(9, 28)=1.65, p=.149)* | | | | | |
|---|---|---|---|---|---|
| Predictor | b | 95% CI | β | t | p |
| Age | -0.02 | [-1.54, 1.51] | -0.00 | -0.02 | .984 |
| Education | -2.78 | [-6.63, 1.07] | -0.30 | -1.48 | .150 |
| Sex | -11.89 | [-33.46, 9.67] | -0.19 | -1.13 | .268 |
| Race | -16.93 | [-39.58, 5.71] | -0.29 | -1.53 | 137 |
| ART delay | 1.81 | [-2.01, 5.64] | 0.24 | 0.97 | .340 |
| Current CD4[1] | -24.60 | [-70.50, 21.30] | -0.20 | -1.10 | .282 |
| Nadir CD4[1] | -13.95 | [-29.70, 1.81] | -0.33 | -1.81 | .080 |
| Disease duration | -0.19 | [-3.37, 2.98] | -0.03 | -0.13 | .901 |
| Viral load[2] | -4.15 | [-31.31, 23.02] | -0.05 | -0.31 | .757 |

| ***BDI-II (total score, n=40)***: *($R^2_{adj}$=.09, F(9, 30)=1.43, p=.220)* | | | | | |
|---|---|---|---|---|---|
| Predictor | b | 95% CI | B | t | p |
| Age | 0.14 | [-0.16, 0.43] | 0.15 | 0.93 | .362 |
| Education | -0.74 | [-1.48, 0.01] | -0.40 | -2.01 | .053 |
| Sex | -3.41 | [-7.60, 0.78] | -0.29 | -1.66 | .106 |
| Race | -4.38 | [-8.72, -0.03] | -0.39 | -2.06 | **.048*** |
| ART delay | 0.53 | [-0.18, 1.25] | 0.36 | 1.52 | .138 |
| Current CD4[1] | -6.07 | [-15.12, 2.99] | -0.25 | -1.37 | .181 |
| Nadir CD4[1] | -0.59 | [-3.69, 2.52] | -0.07 | -0.39 | .703 |
| Disease duration | -0.26 | [-0.87, 0.36] | -0.19 | -0.86 | .399 |
| Viral load[2] | -0.23 | [-5.15, 4.68] | -0.02 | -0.10 | .923 |

b, unstandardized coefficients; β, standardized coefficients;
[1] with log transformed nadir CD4;
[2] binarized viral load (0: <=20 copies/mL; 1: >20 copies/mL); *$p$ < .05

**Supplementary Figure S1. ROC curves for mood and neurocognitive measures in PWH**

Receiver operating characteristic (ROC) curve analysis was performed to evaluate the discriminative ability of neuropsychological measures for distinguishing PWH from control participants. BDI-II (AUC = .712, $^{***}p < .001$) and POMS (AUC = .664, $^{**}p = .002$) showed significant discriminative ability, whereas GMT (AUC = .538, $p$ = .465), and GDS (AUC = .524, $p$ = .643) did not reach statistical significance.

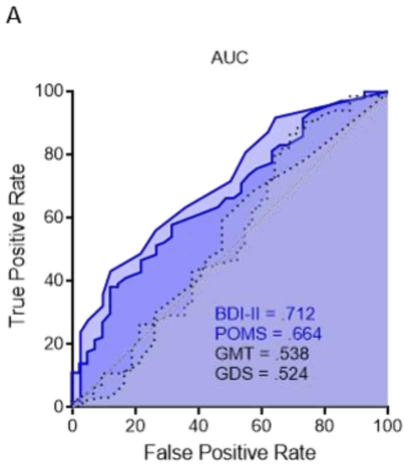

**Supplementary Figure S2. Correlation between mood and neurocognitive measures in PWH**

(A) Total BDI-II scores in PWH showed no significant correlations with GMT ($r$ = -.057, $p$ = .556, $n$ = 109) or GDS ($r$ = .039, $p$ = .684, $n$ = 109).

(B) In contrast, total mood disturbance scores in PWH, as assessed by the POMS, were not significantly correlated with neurocognitive function measures, including GMT ($r$ = -.081, $p$ = .405, $n$ = 107) and GDS ($r$ = .021, $p$ = .827, $n$ = 107).

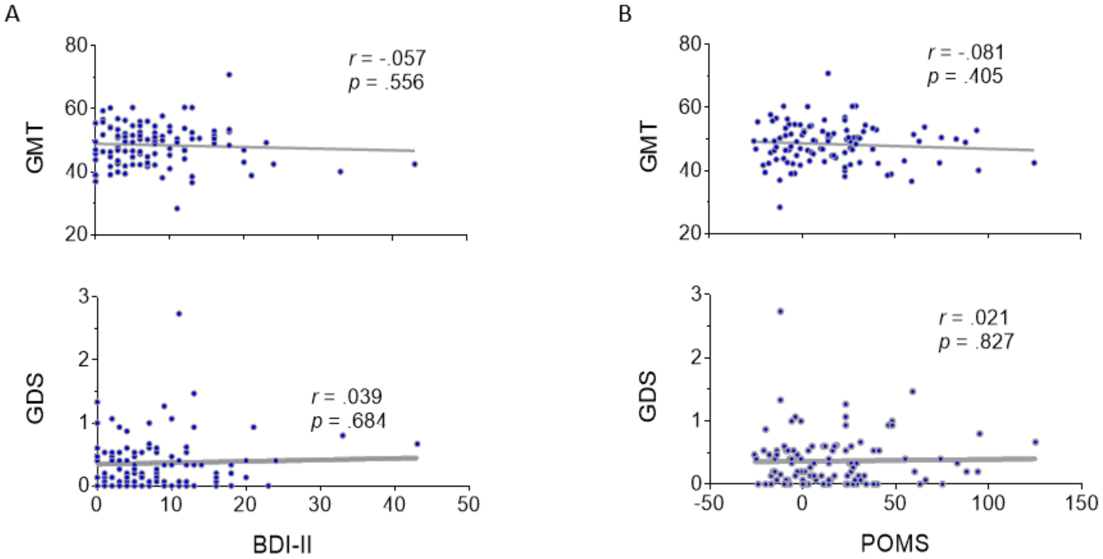

**Supplementary Figure S3. Correlation between duration of ART and mood symptoms in PWH**

(A-B) Duration of ART showed no significant correlations with total BDI-II scores ($r$ = -.068, $p$ = .480, $n$ = 109) and total POMS scores ($r$ = -.111, $p$ = .257, $n$ = 107).

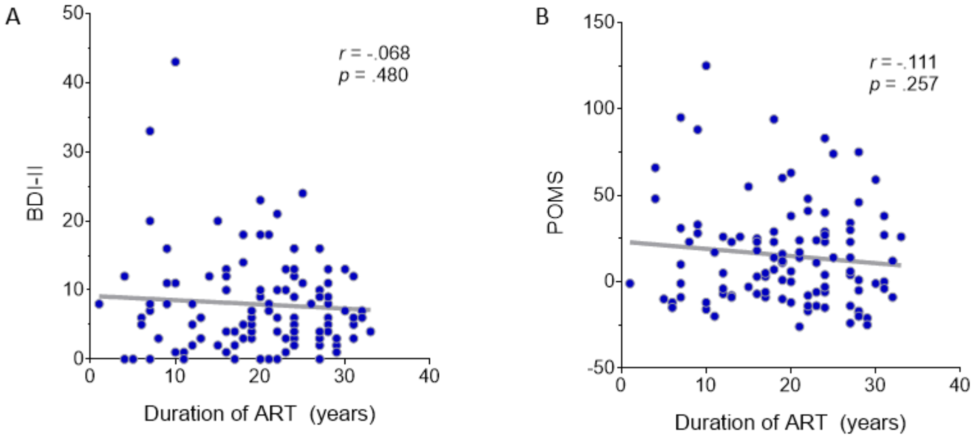
